# Effect of cenegermin (rhNGF) on corneal nerve regeneration and epithelial healing in diabetic neurotrophic keratopathy

**DOI:** 10.1101/2025.05.06.25327131

**Authors:** Rui Wang, Yanling Dong, Qingjun Zhou, Dapeng Sun, Jun Cheng, Qianqian Kong, Xiaochuan Wang, Jiang Bian, Shuang Wang, Bining Zhang, Yangyang Zhang, Lixin Xie

## Abstract

**Introduction:** This study aimed to investigate the impact of cenegermin (recombinant human nerve growth factor) on corneal epithelial healing, nerve regeneration, and tear secretion in patients with diabetic neurotrophic keratopathy(DNK).

**Methods:** This study was conducted as a single-arm and open-label trial. Fourteen patients diagnosed with bilateral DNK were treated with cenegermin eye drops (20 mcg/ml) six times daily for eight weeks and followed up to twenty weeks. Clinical evaluations—including corneal fluorescein staining score, *in vivo* confocal microscopy, corneal sensitivity, and Schirmer I test—were conducted to assess the corneal epithelial healing time, nerve fibre regeneration, and tear secretion.

**Results:** The corneal epithelium exhibited complete healing in week 2, with further enhancement in epithelial integrity observed in week 8 and sustained until week 20. Corneal nerve fibre density (CNFD), length (CNFL), and branch density (CNBD) exhibited a significant increase at week 8 in all patients. However, CNFD and CNFL showed a slight reduction in the peripheral zones of patients with moderate DNK by week 20. Nonetheless, corneal sensitivity demonstrated consistent improvement throughout the treatment period. Tear secretion experienced a significant increase at week 8 but returned to baseline levels by week 20. Furthermore, the best-corrected visual acuity (BCVA) improved after treatment and remained stable until the final visit.

**Conclusions:** The administration of cenegermin effectively facilitated and sustained the healing process of corneal epithelium in cases of mild-to-moderate DNK by promoting nerve regeneration and tear secretion. Prolonged treatment is crucial for stimulating both nerve regeneration and tear secretion.

Trial Registration: Chinese Clinical Trial Registry identifier *ChiCTR2200058806*

**Key Messages:** *What is known?:*

- Diabetic neurotrophic keratopathy (DNK), a subtype of neurotrophic keratopathy (NK) secondary to diabetes mellitus, is characterized by peripheral epithelial defects and varying degrees of corneal hypoesthesia or anesthesia. This condition exhibits poor responsiveness to conventional therapeutic approaches, leading to persistent or progressive lesions.
- The efficacy of rhNGF eye drops (cenegermin) in treating stage 2 and 3 NK has been evaluated in two prospective multicenter randomized clinical trials involving a limited number of patients with moderate and severe DNK. However, these studies do not provide sufficient evidence regarding the effects of rhNGF on tear function, corneal sensitivity and nerve density in mild and moderate DNKs.

*What is new?:*

- The study addresses a significant clinical challenge by investigating an optimized treatment approach for diabetic neurotrophic keratopathy (DNK), focusing on the complex interplay between the corneal nerve, epithelium, and lacrimal gland.
- The administration of cenegermin effectively facilitated and sustained the healing process of the corneal epithelium in cases of mild-to-moderate DNK that were refractory to conventional medical treatments and amniotic membrane transplantation (AMT). Noteworthy findings included significant regeneration of corneal nerve fibers and enhanced tear function.
- Prolonged administration is imperative for promoting nerve regeneration and stimulating tear secretion.

## INTRODUCTION

Corneal sensory nerves originate from the ophthalmic branch of the trigeminal nerve and are responsible for nociception, cold sensation, and pressure perception. It protects the cornea from damage by modulating the blink response, stimulating tear production, and producing trophic factors to maintain homeostasis.[1,2] Neurotrophic factors such as nerve growth factor (NGF) provide trophic support for corneal tissues, stimulate wound healing, and maintain anatomic integrity. Moreover, corneal epithelial cells and keratocytes secrete NGF to facilitate the differentiation and maturation of neurons.[3] This dynamic interplay between neurotrophins, corneal nerves, and corneal epithelium maintains corneal nerve integrity and corneal homeostasis.[4] Consequently, sensory injury to the cornea generally causes impairment of sensory and trophic functions and corneal epithelium breakdown. This affects the health and integrity of the tear film, epithelium, and stroma, clinically defined as neurotrophic keratopathy (NK).[2]

Dua *et al*. proposed a three-stage classification system for NK based on clinical severity and corneal sensitivity: mild, moderate, and severe. Mild NK is characterized by epithelial irregularity without a clear epithelial defect and reduced or absent sensitivity in one or more quadrants of the cornea. Moderate NK is characterized by peripheral epithelial defects (PED) and varying degrees of corneal hypoesthesia or anaesthesia. Severe NK involves stromal ulceration, including frank perforation and corneal hypoesthesia or anaesthesia.[2,5] Diabetic neurotrophic keratopathy (DNK), a form of NK resulting from diabetes, can present as mild (with changes in the epithelium and tear film), moderate (characterized by nonhealing epithelial defects), or severe (involving stromal melting and perforation).[2] In clinical practice, mild DNK is probably more prevalent than moderate and severe forms. Mild DNK is often underestimated and frequently categorized as a refractory dry eye disease; hence, it is not considered for aggressive management.[6] The conventional medical approach, which utilizes lubricants including artificial tears to serum/plasma drops with anti-inflammatory agents, antibiotics, and antiproteases, provides nonspecific relief that may demonstrate transient efficacy.[7] Clinically, the failure of rapid re-epithelialization within 2 weeks of corneal injury usually results in corneal scarring, ulceration, neovascularization, and conjunctivalisation, ultimately causing corneal opacification and vision loss.[8]

Cenegermin 0.002% (20 mcg/ml; recombinant human NGF; Oxervate^®^, Dompè Farmaceutici, Milan, Italy), has been approved in the United States and Europe for treating NK.[9] The efficacy of recombinant human NGF (rhNGF) in treating stage 2 and 3 NK has been investigated in two prospective multicentre randomized clinical trials involving few patients with moderate and severe DNK.[9]^,^[10] These trials demonstrated the effectiveness of rhNGF in promoting corneal healing post-treatment; however, sufficient evidence regarding its impact on corneal sensitivity and nerve density was not provided. Moreover, data regarding the use of cenegermin in patients with stage 1 NK are insufficient. Several retrospective studies have investigated nerve regeneration and tear function in a few mild and moderate cases of DNK, with 4–8 weeks of treatment.[8,11]

The objective of this study was to conduct a prospective trial to evaluate the effects of cenegermin on corneal epithelial wound healing, nerve regeneration, corneal sensitivity, and tear function improvement in patients with mild or moderate DNK who exhibited non-responsiveness to conventional therapy.

## METHODS

### Ethics

This single-arm, single-centre, and open-label trial was approved by the Ethics Committee of Qingdao Eye Hospital at Shandong First Medical University. The study was conducted in accordance with relevant guidelines and adhered to the principles outlined in the Declaration of Helsinki. Written informed consent was obtained from all the participants.

### Study Participants

Participants were recruited between April 15, 2022, and October 15, 2023, and written informed consent was obtained from all individuals.After obtaining written informed consent, 14 patients with type 2 diabetes diagnosed with mild to moderate and bilateral DNK who exhibited non-responsiveness to conventional treatments— such as ocular medications and high-oxygen permeable contact lenses—and amniotic membrane transplantation (AMT) or both were enrolled in this study.[5] Patients enrolled in other clinical trials who had corneal scars that hindered the assessment of corneal nerve changes, experienced active ocular inflammation unrelated to neurotrophic keratitis, and presented with marginal corneal ulcers, herpes simplex keratitis, or a history of corneal refractive surgery or collagen cross-linking surgery were excluded from the study. All enrolled patients demonstrated active management of blood glucose levels, and glycated haemoglobin (HbA1c) was measured at baseline and at weeks 4, 8, and 20 (See Figure S1 in the electronic supplementary material for details).

### Examination and Treatment Procedures

The treatment protocol for 15 eyes with mild DNK involved the administration of 0.002% rhNGF eye drops. Each patient’s eye was treated with one drop four hourly for 8 weeks. In addition to rhNGF eye drops, 13 patients with moderate DNK were administered antibiotics as prophylaxis against ocular infections. All other ocular therapies that could interfere with epithelialization and contact lens bandages were discontinued. Any ocular discomfort or adverse drug reactions experienced by the patients were recorded.

After enrolment, comprehensive systemic and ocular histories of patients were documented. Ocular examinations followed a specific protocol, with a thorough slit-lamp examination and corneal fluorescein staining. This was followed by an assessment of best-corrected visual acuity (BCVA), intraocular pressure (IOP), corneal sensitivity, tear meniscus height (TMH), time to first breakup of the tear film (NIKBUT-first), and average time of all breakup incidents (NIKBUT-average) using Keratograph 5M (K5M; Oculus Optikgerate GmbH, Wetzlar, Germany). Additionally, central corneal thickness was measured using AC-OCT (DRI, Triton, Topcon, Japan), and corneal nerve fibre examination was conducted by corneal confocal microscopy (CCM; Heidelberg Retinal Tomography III with Rostock Cornea Module, Heidelberg Engineering GmbH, Heidelberg, Germany).

Corneal epithelial integrity was evaluated by fluorescein staining using the National Eye Institute (NEI) scale for corneal fluorescein staining, graded from 0 (absent) to 3 (severe) for each of the five areas of the cornea (total score 0–15), with higher scores indicating greater abnormality.[12] Corneal epithelial defects with a maximum diameter <0.5 mm were considered as healing according to the REPARO 2 study.[10]

Corneal sensitivity was assessed using a Cochet–Bonnet esthesiometer (Western Ophthalmics, Lynnwood, Washington, USA) in the central 4 mm zone and the four peripheral sectors (inferior, nasal, superior, and temporal) of the corneal surface. A vertically applied 60 mm nylon filament was gradually decreased in 5-mm increments until a positive response (verbal or blinking) was triggered. The length of the filament at which the response occurred was recorded as the corneal sensitivity threshold. This process was repeated three times, and the average value was obtained.

Corneal nerve fibre parameters were evaluated using the CCM at baseline and at weeks 4, 8, and 20 after treatment. The clinical technician adhered to standardized operating procedures while operating the CCM to sequentially capture digital images at a rate of three frames per second. The “Section” mode was used to record nerve images in the corneal superior, nasal, inferior, temporal, and inferior-whorl zones.

Three non-overlapping scans of the sub-basal nerve plexus (SNP) from five designated regions were analysed using ACCMetrics software (University of Manchester, UK, version 2.0).[13] Quantification was conducted for corneal nerve fibre density (CNFD, the number of fibres/mm^2^), corneal nerve branch density (CNBD, the number of branch points on the main fibres/mm^2^), corneal nerve fibre length (CNFL, the total length of fibres mm/mm^2^), and corneal nerve fibre width (CNFW, average nerve fibre width µm/mm^2^) in the peripheral (p-) and inferior-whorl zones (i-).

### Statistical Analysis

Statistical analysis was performed using IBM SPSS version 25.0 for Mac (IBM Corp., Armonk, NY, USA). Visual acuity was converted to the logarithm of the minimal angle of resolution (logMAR). The Kolmogorov–Smirnov test was used to determine the normal distribution of the quantitative data, presented as the median and interquartile range (IQR). Qualitative variables are presented as frequencies and percentages. The Wilcoxon test was used for pairwise comparisons in each subsequent session. Statistical significance was set at *p*<0.05.

## RESULTS

### Characteristics of Patients

This study included 28 eyes of 14 patients with mild-to-moderate DNK and was conducted between March 21, 2022, and June 17, 2023. Overall, 15 eyes in 11 patients were diagnosed with mild DNK, whereas 13 eyes in 10 patients were diagnosed with moderate DNK. The demographic characteristics and relevant systemic and ocular medical histories of the participants are presented in Table 1. The median (IQR) age of the patients was 58 (16.25) years, with 7 males (50%). The median (IQR) diagnostic course was 11(17.5) years, and all patients had baseline HbA1c levels with a median (IQR) of 6.75% (1.8%).

### Treatment Response

The 8-week treatment course was successfully completed by all patients and followed by a 12-week follow-up period. rhNGF eye drops significantly enhanced the healing of corneal epithelial defects and promoted the restoration of epithelial integrity. After treatment for 2 weeks, all compromised corneal epithelia in moderate DNK cases were completely healed (13/13,100%), with the complete disappearance of punctate staining observed at the end of the treatment period (corneal fluorescein staining, CFS: week 8 vs. week 2, *p*=0.006, Figure 1B, D and Table 3). A similar outcome was also observed in patients with mild DNK (CFS: week 8 vs. baseline, *p*=0.0001, Figure 1 A, C and Table 2).We observed a notable increase in CNFD, CNBD, and CNFL in the corneal peripheral zones at weeks 4 and 8 post-treatment compared with the baseline levels in patients with mild to moderate DNK (Figures 2 and 3 and Tables 2 and 3). Similarly, CNFD and CNFL were increased significantly at weeks 4 and 8 post treatment in the corneal inferior-whorl zones in patients with mild DNK (p=0.034 and p=0.005 at week 4; p=0.034 and p=0.005 at week 8, respectively). Additionally, CNBD was increased significantly only at week 8 post treatment (p=0.018), with no significant difference noted at week 4 (p=0.17) (Figure 2 and Table 2). Corneal neuroinflammation and swelling are the indicators of diabetic corneal neuropathy.[14] CNFW in the corneal inferior whorl zones of mild DNK exhibited a significant decline at weeks 4 (p=0.002) and 8 (p<0.001) post treatment compared with the baseline level. However, no significant differences were observed in the peripheral zones of the mild and moderate DNK groups (Figure 2 and Tables 2 and 3).

**Figure 1.**
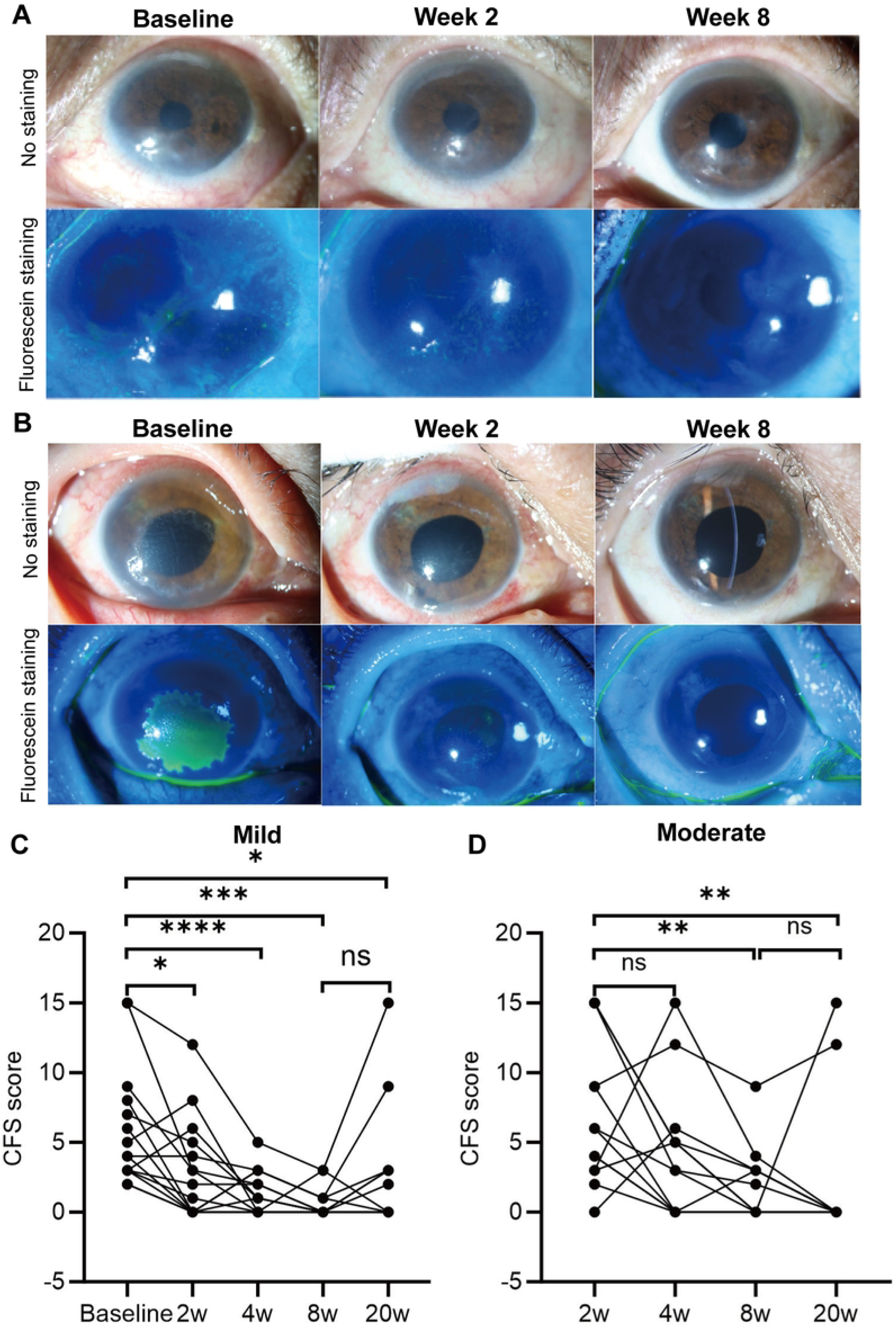
Representative images and CFS score for patients with DNK before and after rhNGF treatment. Anterior segment photos with fluorescein stain shows (A) mild DNK and (B) moderate DNK at baseline, post-treatment week 2, and week 8. Scatter line charts display the differences in CFS score at baseline, and post-treatment weeks 2, 4, 8, and 20 for (C) mild DNK, and post-treatment weeks 2, 4, 8, and 20 for (D) moderate DNK. CFS, corneal fluorescein staining; DNK, diabetic neurotrophic keratopathy; ns, no significant difference; rhNGF, recombinant human nerve growth factor. \**p*<0.05, \*\**p*<0.01, \*\*\**p*<0.005 and \*\*\*\**p*<0.0001.

**Figure 2.**
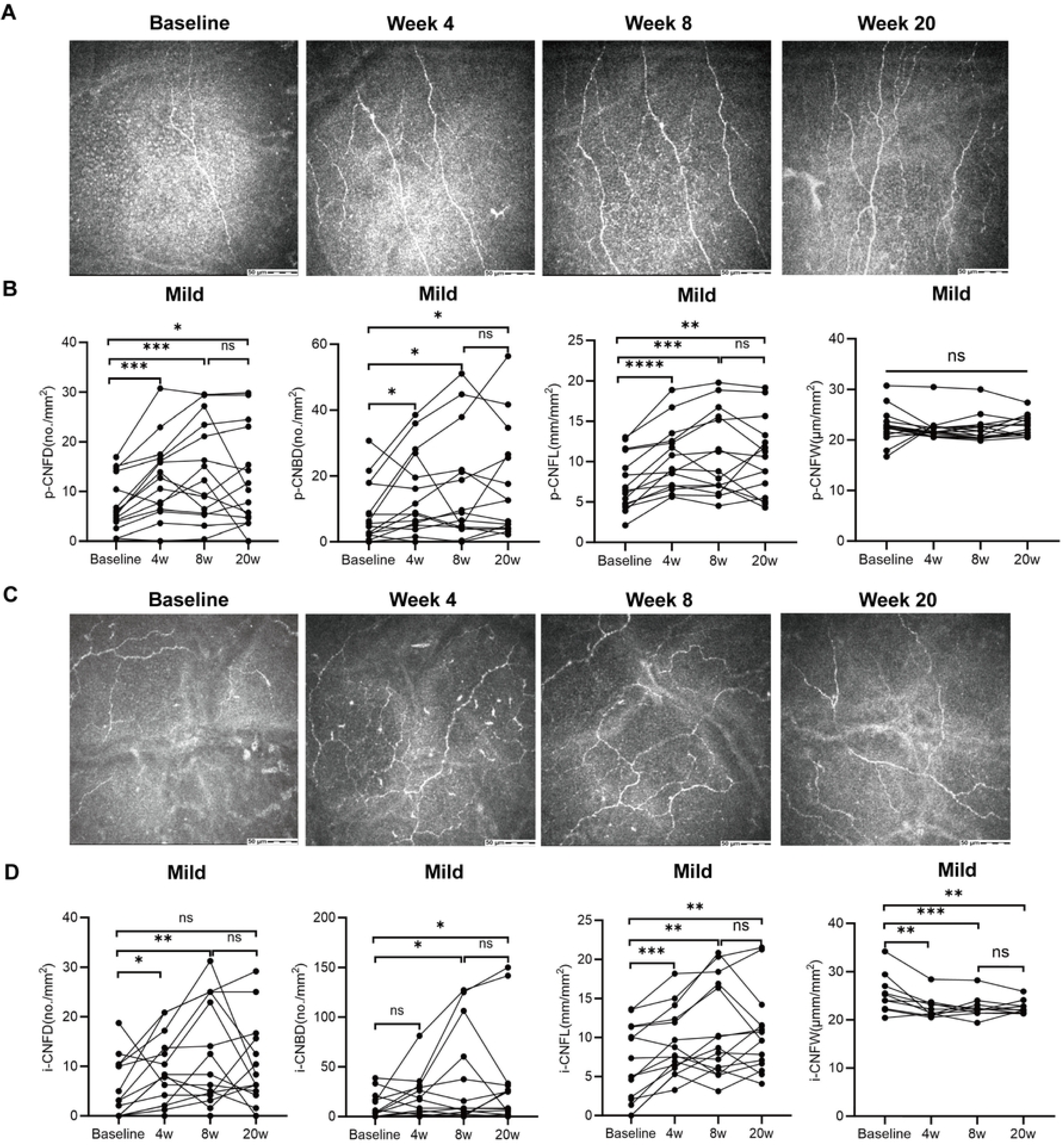
Representative CCM images and nerve fibre parameters in patients with mild DNK before and after rhNGF treatment. CCM images of the (A) corneal peripheral zone and (C) inferior-whorl zone are observed at baseline, and post-treatment weeks 4, 8, and 20 in mild DNKs. Scatter line charts display (B) the differences in p-CNFD, p-CNBD, p-CNFL, and p-CNFW of the corneal peripheral zone at baseline, and post-treatment weeks 4, 8, and 20, and (D) the variances in i-CNFD, i-CNBD, i-CNFL, and i-CNFW of the corneal inferior-whorl at baseline, and post-treatment weeks 4, 8, and 20. CCM, corneal confocal microscopy; CNBD, corneal nerve branch density; CNFD, corneal nerve fibre density; CNFL, corneal nerve fibre length; CNFW, corneal nerve fibre width; DNK, diabetic neurotrophic keratopathy; rhNGF, recombinant human nerve growth factor; ns, no significant difference; \**p*<0.05, \*\**p*<0.01, \*\*\**p*<0.005 and \*\*\*\**p*<0.0001.

**Figure 3.**
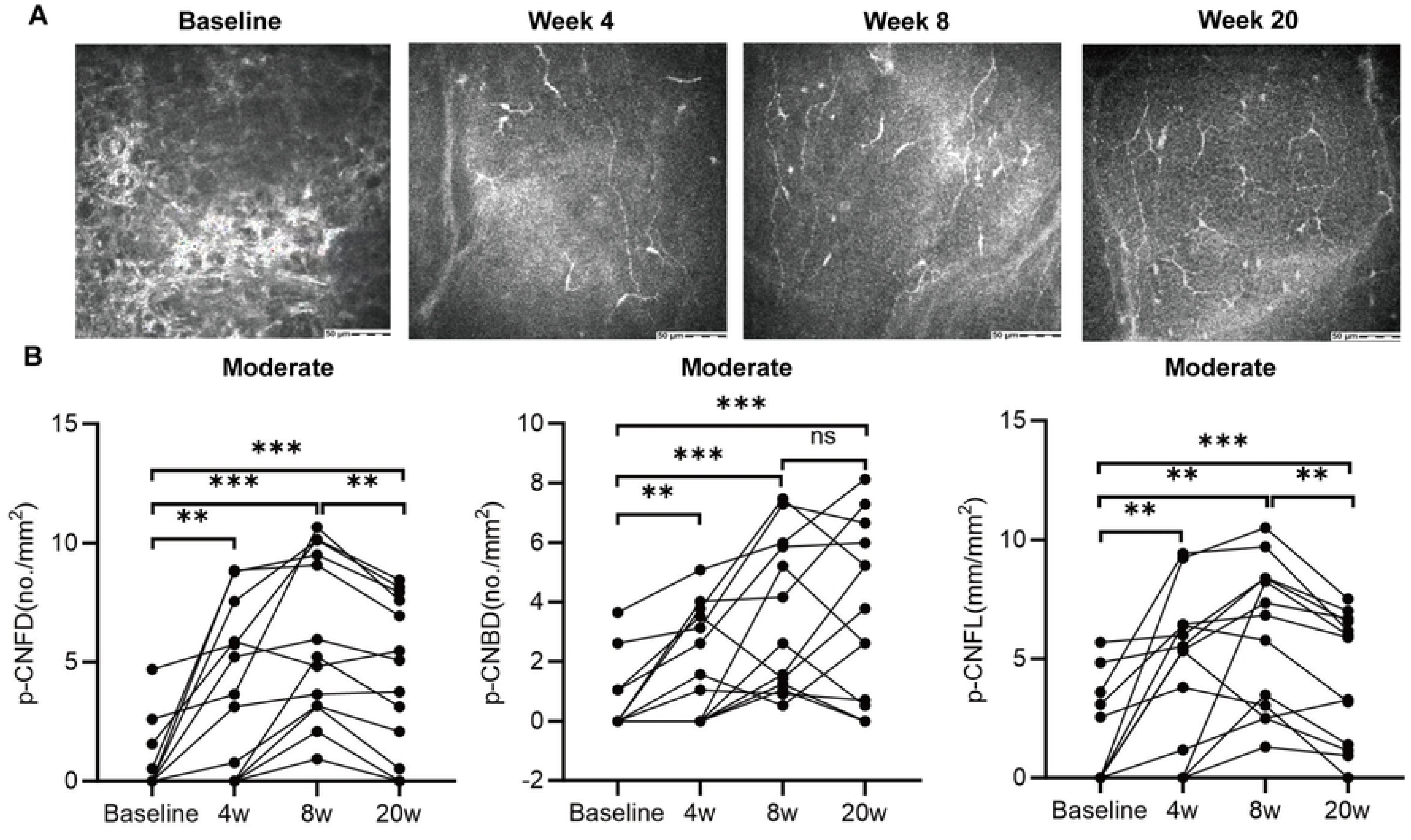
Representative CCM images and nerve fibre parameters are assessed for patients with moderate DNK before and after rhNGF treatment. (A) CCM images of the corneal peripheral zone are obtained at baseline, and post-treatment weeks 4, 8, and 20 in mild DNKs. (B) Scatter line charts are used to illustrate the differences in p-CNFD, p-CNBD, and p-CNFL of the corneal peripheral zone at baseline, and post-treatment weeks 4, 8, and 20. CCM, corneal confocal microscopy; CNBD, corneal nerve branch density; CNFD, corneal nerve fibre density; CNFL, corneal nerve fibre length; CNFW, corneal nerve fibre width; DNK, diabetic neurotrophic keratopathy; rhNGF, recombinant human nerve growth factor; ns: no significant difference; w, week. * *p*<0.05, \*\**p*<0.01 and \*\*\**p*<0.005.

Corneal sensitivity and tear production increased at weeks 2, 4, and 8 post treatment in patients with mild DNK and at weeks 4 and 8 in those with moderate DNK (Tables 2 and 3 and Figure 4). However, no significant changes were observed in the TMH, NIKBUT-first, and average following treatment compared to the baseline levels in patients with mild or moderate keratopathy, except for a slight increase in NIKBUT-first at post-treatment week 8 in patients with moderate keratopathy (Tables 2 and 3).

**Figure 4.**
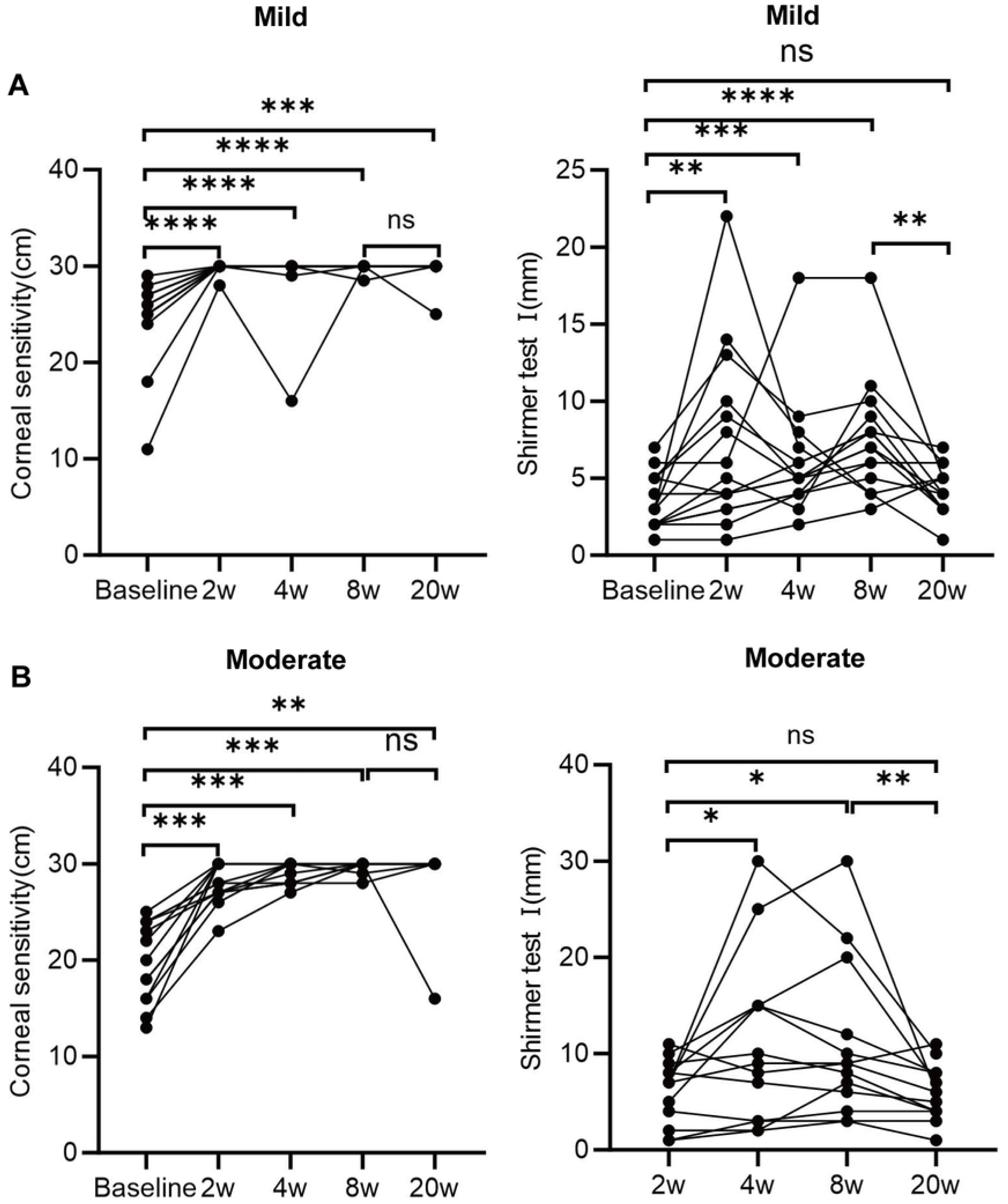
Differences in corneal sensitivity and tear secretion function in patients with mild and moderate DNK are assessed at baseline and after rhNGF treatment. The scatter line charts illustrate variations in corneal sensitivity and Shirmer test I among (A) mild DNKs at baseline, and post-treatment weeks 2, 4, 8, and 20, and among moderate DNKs at baseline, and post-treatment weeks 2, 4, 8, and 20 for corneal sensitivity and at post-treatment weeks 2, 4, 8, and 20 for Schirmer test I. DNK, diabetic neurotrophic keratopathy; rhNGF, recombinant human nerve growth factor; ns: no significant difference; w, week. \**p*<0.05, \*\**p*<0.01, \*\*\**p*<0.005 and \*\*\*\**p*<0.0001.

The BCVA is used to assess efficacy outcomes. We observed a slight increase in mild keratopathy at the end of treatment (*p*=0.007). In contrast, patients with moderate keratopathy exhibited significantly higher levels following treatment than at baseline (*p*=0.007, *p*=0.029, *p*=0.002 at weeks 2, 4, and 8, respectively). This result can be attributed to a significant thinning of the central corneal thickness to the normal range after treatment with rhNGF eye drops in patients with mild (*p*=0.028 and *p*=0.005 at weeks 4 and 8, respectively) and moderate keratopathy (*p*=0.021 and *p*=0.008 at weeks 4 and 8, respectively) (Tables 2 and 3, and Figure 5).

**Figure 5.**
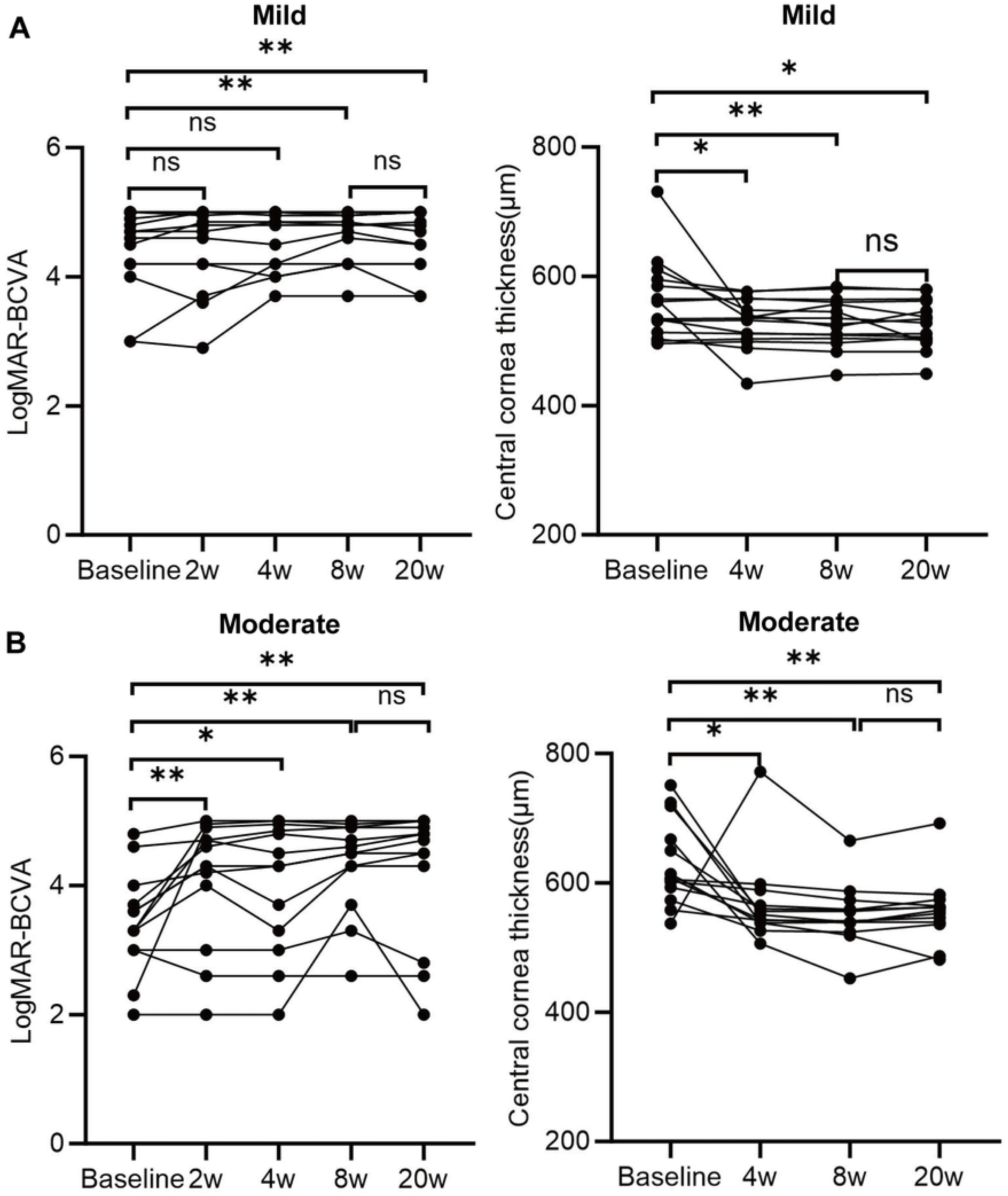
Differences in BCVA and central corneal thickness in patients with mild and moderate DNK are assessed at baseline and after rhNGF treatment. The scatter line charts show the changes in BCVA and central corneal thickness among (A) mild and (B) moderate DNKs at baseline, and post-treatment weeks 4, 8, and 20. BCVA, best corrected visual acuity; ns: no significant difference; w, week. \**p*<0.05 and \*\**p*<0.01.

The long-term efficacy of rhNGF eye drops was observed during the follow-up visit, demonstrating a significant improvement compared with the baseline values. rhNGF preserved corneal epithelial integrity until the end of the follow-up period (*p*>0.05), compared with data at week 8 (Tables 2 and 3). Importantly, no differences were observed in CNFD, CNBD, CNFL, and CNFW in the peripheral zones and inferior whorl zone compared with data at week 8; moreover, these parameters showed consistent improvement from baseline levels in mild DNK cases (Figure 2). However, in moderate DNK cases, a significant reduction was noted in CNFD and CNFL in the peripheral zones at week 20 (*p*=0.001 and *p*=0.001, respectively; Figure 3); nevertheless, these values remained significantly higher than baseline levels (*p*=0.001 and *p*=0.005, respectively; Figure 3). Notably, tear secretion function decreased at week 20 post treatment compared with week 8 (*p*=0.03 and *p*=0.005), with no statistically significant difference compared with the baseline level in mild DNK (*p*=0.192) or week 2 level in moderate DNK (*p*=0.906) (Figure 4).

rhNGF improved corneal sensitivity until 12 weeks post treatment in patients with mild (*p*>0.999 compared with week 8 and *p*=0.0001 compared with baseline) and moderate (*p*>0.999 compared with week 8 and *p*=0.003 compared with baseline) DNK, despite a slight decrease in CNFD and CNFL (Tables 2 and 3; Figure 4). At the end of the visit, BCVA and central corneal thickness significantly improved for all patients compared with that at baseline (*p*=0.031 and *p*=0.007, respectively, for BCVA; *p*=0.014 and *p*=0.008, respectively, for central corneal thickness; Tables 2 and 3; Figure 5).

### Safety

There were no instances of treatment discontinuation due to drug-related adverse effects. The adverse events observed in the patients were mild, transient, and well tolerated, including eye irritation (2/24, 16.7%) and burning sensations (4/24, 33.3%). In addition, no difference in IOP was observed between baseline and follow-up visits.

## DISCUSSION

Corneal epitheliopathy is characterized by superficial punctate keratitis, recurrent epithelial erosion, persistent epithelial defects, and delayed (often incomplete) wound healing.[5] Corneal epithelial wound healing plays a vital role in restoring corneal barrier function after injury. However, in patients with diabetes, corneal epithelial damage is often accompanied by neuropathy and alterations in tear film composition, resulting in prolonged healing times or even non-healing, thereby presenting challenges for the effective treatment of diabetic corneal erosion.[7] Healthy corneas rely on proper functioning and communication between epithelial cells and sensory neurons, and diabetes mellitus (DM) perturbs the interaction and interdependencies between these two cells.[15]

Reportedly, NGF plays a key role in the modulation of immune reactions, trophic support, ocular surface healing, corneal sensitivity, and tear film function.[16,17] This study presents a prospective cohort investigation, demonstrating the rapid and effective clinical efficacy and long-term maintenance of rhNGF in patients with mild-to-moderately refractory DNK. Within a 2-week treatment period, all patients exhibited complete healing of the corneal epithelial defects. Furthermore, significant improvements were observed in CNFD and CNFL, corneal sensitivity, tear secretion, and BCVA after treatment for 8 weeks. Importantly, moderate DNK cases showed partial regression in CNFD and CNFL, whereas tear secretion function returned to baseline levels across all DNK cases during the 12-week follow-up period.

In our study, all patients with DNK showed significant improvement in corneal epithelial integrity, particularly those with moderate DNK, who experienced rapid healing within 2 weeks despite imperceptible regeneration of corneal nerves. These results are consistent with findings from the two largest clinical trials that investigated rhNGF.[8–10] The development of corneal epithelial lesions in patients with diabetes is attributed to neurodegeneration, decreased expression of NGFs, impaired tear film function, accumulation of glycosylation end products, and heightened oxidative stress.[7] Exogenous NGF stimulates corneal nerve regeneration and directly affects corneal epithelial cells, thereby promoting corneal epithelial healing.[18,19] The entire patient cohort comprised individuals who did not respond to multiple standard therapies. In our study, 6 of the 28 eyes underwent AMT treatments, while 2 received three rounds of AMTs; however, no positive response was observed. Notably, all patients with DNK exhibited complete corneal epithelial healing and improved vision following rhNGF treatment, which persisted until the end of the visit. Therefore, the use of rhNGF eye drops as a management strategy holds potential for generalization in mild and moderate DNK, where multiple standard therapies prove ineffective.

The present study observed an improvement in sub-basal nerve regrowth after treatment in patients with DNK, which is consistent with previous findings.[20,21] Corneal nerve density and nerve length were increased at 4 weeks, after 8 weeks, and during the follow-up assessments up to 12 weeks post-therapy in the mild and moderate DNKs groups. Substantial centripetal growth of corneal nerves was not observed after 8 weeks of treatment in individuals with moderate DNK (See Figure S2 in the electronic supplementary material for details). This group also displayed limited nerve regeneration in the peripheral zones, which aligns with the findings reported by Pedrotti *et al.*[22] Maintenance of corneal surface homeostasis depends on the interaction of multiple factors, including the role of the corneal epithelium and nerves.[2] The corneal nerves release trophic factors that regulate the proliferation, differentiation, and integrity of corneal epithelial cells. In contrast, the corneal epithelium promotes nerve growth, maturation, and survival. Any disruption of these components compromises the equilibrium of the corneal surface.[3,23] Notably, there was a slight degeneration of corneal nerve fibres in the peripheral zones of moderate rather than mild DNKs after discontinuation for 12 weeks, contradicting the findings of a previous study conducted on stage 2 and 3 NKs.[22] This finding suggests that a gradual decrease in endogenous NGF could occur, if early nerve restoration in patients with diabetes with a deteriorating corneal microenvironment causing decreased NGF secretion by corneal epithelial cells is unsuccessful. This can disrupt corneal surface homeostasis due to insufficient long-term production. Therefore, in patients with mild or severe DNK, continuous administration of NGF should be maintained until a balance between its secretion and consumption by regenerating nerves and the corneal epithelium is achieved.

Corneal sensitivity was rapidly recovered, although the regrowth of sub-basal nerve fibres was less noticeable in moderate DNKs after 8 weeks of treatment and during the follow-up evaluations for up to 12 weeks post therapy, suggesting effective sensory transmission. Corneal sensitivity recovers faster than objective nerve detection after epithelium-off crosslinking and LASIK procedures,[24] which is likely due to the limitations of slit-scanning confocal microscopy in detecting fine regenerating nerves rather than their actual absence. Diode laser-equipped *in vivo* confocal microscopy provides higher contrast and a shallower step size compared to slit scanning confocal microscopy, enabling better identification of subtle nerves.[25] The prevalence of dry eye in diabetes has been reported to be as high as 54.3%,[26] which is more commonly observed than corneal symptoms among patients with diabetes.[27] Our study is the first to assess the efficacy of rhNGF in ameliorating the clinical manifestations of dry eye syndrome in patients with DNK and demonstrated that tear secretion ability is rapidly improved following topical application of NGF eye drops, corresponding to a recent phase II clinical trial assessing the role of rhNGF in patients with dry eye diseases (DED), which also showed significant improvements in DED signs and symptoms after 4 weeks of treatment.[28] Tear secretion is regulated by the afferent sensory nerves originating from the cornea and the efferent autonomic nerves that innervate the lacrimal and meibomian glands.[17,29] Corneal denervation in diabetes leads to impaired afferent signalling and reduced neurotransmitters, leading to reduced tear production and DED.[29,30] NGF has been shown to stimulate tear film production and functionality *in vitro* and *in vivo*, providing potential benefits to patients with DED.[31] However, the promotional effect of NGF on tear secretion was not sustained beyond 12 weeks after withdrawal, as it returned to pre-treatment levels. In a previous study, the positive function of rhNGF was extended until 4 weeks after discontinuation in patients with moderate-to-severe DED and non-diabetics.[28] Sacchetti *et al*. clarified that the therapeutic effect of rhNGF eye drops on DED is dose-dependent, with significantly greater efficacy observed at higher concentrations than at lower concentrations.[28] The pathogenesis of dry eye syndrome in diabetes involves the dysfunction of the lacrimal glands, meibomian glands, conjunctival goblet cells, corneal epithelial cells, and nerve fibres. These dysfunctions cause alterations in the quantity and quality of tears, leading to tear film instability.[32–35] Therefore, future clinical trials should explore the appropriate concentration and duration of rhNGF eye drops to manage DNK effectively.

Inadequate and ineffective management of diabetic keratopathy often results in compromised visual acuity or permanent vision loss.[5] Corneal oedema is a prominent clinical feature in patients with DNK.[5] In our study, BCVA exhibited a significant increase, accompanied by a notable reduction in central corneal thickness after rhNGF treatment and the final visit, indicating enhanced corneal transparency. A retrospective study including 17 patients diagnosed with stage 1 NK also demonstrated a significant improvement in BCVA after treatment with rhNGF.[21] In contrast, the BCVA may not improve significantly because of corneal thinning and/or subepithelial scarring in severe DNK. Therefore, early treatment of NK (mild or moderate) is important to retain BCVA and prevent permanent loss of vision.[21,36,37] Restoration of corneal transparency after wound healing is a multifaceted process requiring an integrated response. Neurotrophic factors secreted by the sensory nerves can promote corneal wound healing and improve corneal transparency by suppressing TGF-1-mediated tissue fibrosis, decreasing neutrophil infiltration and macrophage activation, inhibiting the expression of pro-inflammatory mediators, and preserving corneal endothelial cell density and function.[38]

The primary strength of this prospective cohort study lies in its comprehensive reporting of the progressive impact of topical rhNGF on epithelial wound healing, nerve regeneration, improved corneal sensitivity, and tear secretion in patients with mild-to-moderate DNK. These findings suggest that for patients with mild-to-moderate DNK, the treatment duration can be shortened or transitioned to a lower concentration of rhNGF after corneal epithelium healing. However, achieving full improvement in tear secretion in patients with diabetes requires prolonged and sustained administration of rhNGF.

Nevertheless, the present study has certain limitations. First, the absence of a control group was due to the unavailability of a suitable comparator treatment. Additionally, the recruitment of patients with severe DNK was impeded by the COVID-19 pandemic and the discontinuation of rhNGF in China. Consequently, the observations of patients with severe DNKs could not be performed. There are also limitations pertaining to the small sample size and short follow-up duration.

## CONCLUSION

This study demonstrated the high efficacy of topical rhNGF eye drops for mild to moderate DNK in a small cohort and validated their role in improving tear secretion function in patients with diabetes. These findings provide preliminary support for the clinical application of treatments for diabetic dry eye and highlight the need for future investigations to determine the optimal dosage and duration of therapy required to achieve a sustained therapeutic effect in patients with DNK.

## Data Availability

All relevant data are within the manuscript and its Supporting Information files.

## Author Contribution

Rui Wang designed the study. Rui Wang, Yanling Dong, Dapeng Sun, Jun Cheng, Qianqian Kong, Xiaochuan Wang, Jiang Bian and Shuang Wang collected the data. Qingjun Zhou and Bining Zhang helped check the data. Yangyang Zhang,and Lixin Xie contributed to the interpretation of the results and critical revision of the manuscript. All the authors have read and approved the final manuscript. Lixin Xie and Yangyang Zhang are the study guarantors.

## Data Availability

The datasets generated during and/or analyzed during the current study are available from the corresponding author on reasonable request.

## Declarations

### Ethical Approval

The study was approved by the Ethics Committee of Qingdao Eye Hospital at Shandong First Medical University (2021(01)) and was conducted in compliance with the principles of the Declaration of Helsinki. Written informed consent was obtained from all patients.

## REFERENCES

1. You, L., F.E. Kruse, and H.E. Volcker, Neurotrophic factors in the human cornea. Invest Ophthalmol Vis Sci, 2000. 41(3): p. 692–702.

2. Dua, H.S., et al., Neurotrophic keratopathy. Prog Retin Eye Res, 2018. 66: p. 107–131.

3. Di, G., et al., Corneal Epithelium-Derived Neurotrophic Factors Promote Nerve Regeneration. Invest Ophthalmol Vis Sci, 2017. 58(11): p. 4695–4702.

4. Al-Aqaba, M.A., et al., Corneal nerves in health and disease. Prog Retin Eye Res, 2019. 73: p. 100762.

5. Yu, F.X., et al., The impact of sensory neuropathy and inflammation on epithelial wound healing in diabetic corneas. Prog Retin Eye Res, 2022. 89: p. 101039.

6. Hsu, H.Y. and D. Modi, Etiologies, Quantitative Hypoesthesia, and Clinical Outcomes of Neurotrophic Keratopathy. Eye Contact Lens, 2015. 41(5): p. 314–7.

7. Priyadarsini, S., et al., Diabetic keratopathy: Insights and challenges. Surv Ophthalmol, 2020. 65(5): p. 513–529.

8. Dai, X., et al., Effect of Topical Recombinant Human Nerve Growth Factor on Corneal Epithelial Regeneration in Refractory Epithelial Keratopathy. Ocul Immunol Inflamm, 2024: p. 1–7.

9. Pflugfelder, S.C., et al., Topical Recombinant Human Nerve Growth Factor (Cenegermin) for Neurotrophic Keratopathy: A Multicenter Randomized Vehicle-Controlled Pivotal Trial. Ophthalmology, 2020. 127(1): p. 14–26.

10. Bonini, S., et al., Phase II Randomized, Double-Masked, Vehicle-Controlled Trial of Recombinant Human Nerve Growth Factor for Neurotrophic Keratitis. Ophthalmology, 2018. 125(9): p. 1332–1343.

11. Balbuena-Pareja, A., et al., Effect of recombinant human nerve growth factor treatment on corneal nerve regeneration in patients with neurotrophic keratopathy. Front Neurosci, 2023. 17: p. 1210179.

12. Lemp, M.A., Report of the National Eye Institute/Industry workshop on Clinical Trials in Dry Eyes. CLAO J, 1995. 21(4): p. 221–32.

13. Williams, B.M., et al., An artificial intelligence-based deep learning algorithm for the diagnosis of diabetic neuropathy using corneal confocal microscopy: a development and validation study. Diabetologia, 2020. 63(2): p. 419–430.

14. Roszkowska, A.M., et al., Corneal nerves in diabetes-The role of the in vivo corneal confocal microscopy of the subbasal nerve plexus in the assessment of peripheral small fiber neuropathy. Surv Ophthalmol, 2021. 66(3): p. 493–513.

15. Gao, N., et al., Dendritic cell dysfunction and diabetic sensory neuropathy in the cornea. J Clin Invest, 2016. 126(5): p. 1998–2011.

16. Lambiase, A., M. Sacchetti, and S. Bonini, Nerve growth factor therapy for corneal disease. Curr Opin Ophthalmol, 2012. 23(4): p. 296–302.

17. Labetoulle, M., et al., Role of corneal nerves in ocular surface homeostasis and disease. Acta Ophthalmol, 2019. 97(2): p. 137–145.

18. Blanco-Mezquita, T., et al., Nerve growth factor promotes corneal epithelial migration by enhancing expression of matrix metalloprotease-9. Invest Ophthalmol Vis Sci, 2013. 54(6): p. 3880–90.

19. Lambiase, A., et al., Nerve growth factor promotes corneal healing: structural, biochemical, and molecular analyses of rat and human corneas. Invest Ophthalmol Vis Sci, 2000. 41(5): p. 1063–9.

20. Mastropasqua, L., et al., In Vivo Evaluation of Corneal Nerves and Epithelial Healing After Treatment With Recombinant Nerve Growth Factor for Neurotrophic Keratopathy. Am J Ophthalmol, 2020. 217: p. 278–286.

21. Yavuz Saricay, L., et al., Efficacy of Recombinant Human Nerve Growth Factor in Stage 1 Neurotrophic Keratopathy. Ophthalmology, 2022. 129(12): p. 1448–1450.

22. Pedrotti, E., et al., Eight months follow-up of corneal nerves and sensitivity after treatment with cenegermin for neurotrophic keratopathy. Orphanet J Rare Dis, 2022. 17(1): p. 63.

23. Sacchetti, M. and A. Lambiase, Neurotrophic factors and corneal nerve regeneration. Neural Regen Res, 2017. 12(8): p. 1220–1224.

24. Cruzat, A., Y. Qazi, and P. Hamrah, In Vivo Confocal Microscopy of Corneal Nerves in Health and Disease. Ocul Surf, 2017. 15(1): p. 15–47.

25. Patel, D.V. and C.N. McGhee, In vivo confocal microscopy of human corneal nerves in health, in ocular and systemic disease, and following corneal surgery: a review. Br J Ophthalmol, 2009. 93(7): p. 853–60.

26. Achtsidis, V., et al., Dry eye syndrome in subjects with diabetes and association with neuropathy. Diabetes Care, 2014. 37(10): p. e210–1.

27. Zou, X., et al., Prevalence and clinical characteristics of dry eye disease in community-based type 2 diabetic patients: the Beixinjing eye study. BMC Ophthalmol, 2018. 18(1): p. 117.

28. Sacchetti, M., et al., Effect of recombinant human nerve growth factor eye drops in patients with dry eye: a phase IIa, open label, multiple-dose study. Br J Ophthalmol, 2020. 104(1): p. 127–135.

29. Ma, L., et al., CGRP Released by Corneal Sensory Nerve Maintains Tear Secretion of the Lacrimal Gland. Invest Ophthalmol Vis Sci, 2024. 65(4): p. 30.

30. Misra, S.L., et al., Peripheral neuropathy and tear film dysfunction in type 1 diabetes mellitus. J Diabetes Res, 2014. 2014: p. 848659.

31. Coco, G., et al., Cenegermin for the treatment of dry eye disease. Drugs Today (Barc), 2023. 59(3): p. 113–123.

32. Zhang, S., et al., Hyperglycemia Induces Tear Reduction and Dry Eye in Diabetic Mice through the Norepinephrine-alpha(1) Adrenergic Receptor-Mitochondrial Impairment Axis of Lacrimal Gland. Am J Pathol, 2023. 193(7): p. 913–926.

33. Ljubimov, A.V. and M. Saghizadeh, Progress in corneal wound healing. Prog Retin Eye Res, 2015. 49: p. 17–45.

34. Wang, H., et al., Lipidomic analysis of meibomian glands from type-1 diabetes mouse model and preliminary studies of potential mechanism. Exp Eye Res, 2021. 210: p. 108710.

35. Weng, J., et al., Diabetes-Associated Hyperglycemia Causes Rapid-Onset Ocular Surface Damage. Invest Ophthalmol Vis Sci, 2023. 64(14): p. 11.

36. Qu, Y., et al., A New Treatment for Recalcitrant Neurotrophic Keratopathy of Ocular Graft-Versus-Host Disease with Virus Infection. Ophthalmol Ther, 2024. 13(2): p. 469–479.

37. Roszkowska, A.M., et al., Clinical and instrumental assessment of the corneal healing in moderate and severe neurotrophic keratopathy treated with rh-NGF (Cenegermin). Eur J Ophthalmol, 2022. 32(6): p. 3402–3410.

38. Zidan, A.A., et al., Topical application of calcitonin gene-related peptide as a regenerative, antifibrotic, and immunomodulatory therapy for corneal injury. Res Sq, 2023.

